# *‘Giving birth is like going to war’:* Obstetric violence in public maternity centers in Niger

**DOI:** 10.1101/2023.06.26.23291780

**Authors:** Amina P. Alio, Rahmatou M. Garba, Mona Mittal, Anna P. McCormick, Moha Mahamane

**Affiliations:** Department of Public Health Sciences, University of Rochester Medical Center, Rochester, NY; Faculté des Sciences Médicales, Université Abdou Moumouni, Niamey, Niger; Department of Family Science, University of Maryland, College Park, NY; Laboratoire d’Etudes et de Recherches sur les Dynamiques Sociales et le Développement Local (LASDEL), Niamey, Niger

## Abstract

Obstetric violence has been recognized as a significant risk factor for maternal morbidity and mortality globally. However, there is lack of literature on the abuse of birthing women and their maternal rights in formal healthcare settings in Niger. This paper explores women’s experiences of violence in public maternity care in Niger to identify drivers, facilitators, manifestations, and consequences of the maltreatment of women during labor and delivery. We qualitatively explored the experiences of women who gave birth in a public maternity hospital or clinic within the 5 years preceding the study. We conducted five key informant interviews to help inform recruitment and interview protocols, a listening session with eight mothers, and individual interviews with 50 women from four areas of the urban region of Niamey. The 58 participating mothers shared experiences that included physical abuse (slapping), verbal abuse (insults), psychological abuse (mockery), discrimination (social status/ economic status), financial coercion (demanding money prior to care), and harmful birthing practices (forcing the baby out with elbows). Socio-cultural aspects driving maltreatment of women included beliefs about expression of pain during labor and delivery, while social connection and the presence of the physician during delivery were protective factors. Our study findings reflect global concerns about obstetric violence and its consequences. Given the high burden of maternal and infant morbidity and mortality in Niger, it is critical to train clinical staff in safe and respectful maternity care, to improve supervision of care, to institute legal recourse for women, and to consider the integration of traditional birth attendants in clinical settings.

## Introduction

Globally every year women experience neglectful, rude, abusive, or violent treatment during pregnancy, childbirth, and post-partum. These experiences have been conceptualized as obstetric violence (1). Obstetric violence is a form of reproductive violence at the intersection of institutional/structural violence and gender-based violence. Obstetric violence encompasses all forms of harm and maltreatment during pregnancy, delivery, and the post-partum period. However, at the core of the phenomenon is a violation of bioethical principles by health care professionals, the health facility, health system, or other society governing systems that results in women being harmed (2). Obstetric violence has been identified as a barrier to reducing maternal morbidity and mortality globally (3,4).

In 2010, Bowser and Hill catalyzed international efforts to define, document, and measure obstetric violence (5). They proposed seven categories of violent experiences– “physical abuse, non-consented clinical care, non-confidential care, non-dignified care, discrimination, abandonment, and detention in health facilities” (5). These categories were later adopted by the WHO Respectful Maternal Care Charter with some modifications (6). More recently these categories were refined to include: “physical abuse, sexual abuse, verbal abuse, stigma and discrimination, failure to meet professional standards of care, poor rapport between women and providers, and health system conditions and constraints” (7). Examples of physical obstetric violence include being slapped, pinched, beaten, restrained or tied down during labor, and also unconsented medically examination or forced medical procedures such as shaving of pubic hair and episiotomy without one’s consent (8–10). Sexual abuse during childbirth involves sexual harassment, rape, and other sexual violations by health professionals and other medical staff (8). Experiences of verbal abuse include being shouted at, scolded, called derogatory names, threatened with withholding of treatment, singled out with negative or discriminatory comments (7,11). A few studies have documented the failure to meet professional standards of care. For example, lack of privacy during childbirth has been found to be severe and dehumanizing, including, being forced to be on all fours, unclothed and totally exposed while being observed by large groups of students or multiple health attendants (12–15). Extant literature points to medical professionals and staff across medical health systems as the primary perpetrators of obstetric violence (3,7,11).

The complex nature of obstetric violence creates challenges for accurate measurement of incidents of obstetric violence. Currently, there are no global estimates of how many women have experienced abuse or harm during childbirth in their lifetime. There are a handful of studies that document the prevalence of obstetric violence in health facilities in some countries. For example, in Nigeria 35% of mothers interviewed at 6-weeks postpartum during a child immunization clinic visit reported experiencing physical abuse during labor and delivery, with 17% reporting being restrained or tied down during labor (10). More than 25% of the women interviewed for this study also reported episiotomy without consent (10). Studies also show that neglect and abandonment during childbirth is a frequent experience. Fifteen and a half percent of Tanzanian women and 29% of Nigerian women reported neglect and abandonment during childbirth and 3.9-5.4% respectively reported that they delivered alone (8,10). In Tanzania, 13.2% of women self-reported experiencing verbal abuse such as being shouted at, scolded, called stupid, or threatened (8). Alarmingly, in a study conducted in Tanzania, 81.6% of the women directly observed during labor and delivery were not asked for consent for vaginal examination in the antenatal ward, 91.5% of the women were detained in the health facility, and 62.7% of the women were not provided with a clean bed in the postnatal ward (9). Numerous qualitative studies have recorded women’s report of these same experiences in countries around the world (7,11).

A small body of literature shows that women report experiences of discrimination and obstetric violence during childbirth based on their ethnicity, race, faith tradition, age- being an adolescent or being older, relationship status, presence of partner, parity, HIV-status, and socioeconomic status (3,10,16). Other risk factors that have been identified include: level of education, residence in rural or impoverished areas, or being perceived to be unclean (3,8–11). In addition to individual risk factors, many studies link health system and structural factors such as dilapidated buildings, old medical equipment, staffing shortages, improperly or poorly trained staff, unsupervised staff, and lack of supplies to women’s experience of obstetric abuse (7–11). Health system mismanagement and inadequate systems of accountability may lead to staffing shortage and situations in which women can be bribed or extorted or compelled to pay medical fees or bribes in order to receive services (11,17–19). Other types of maltreatment by staff include unreasonable requests such as reports of being forced to clean up blood or other bodily fluids immediately after vaginal and cesarean childbirth (11).

In Niger, out of every 100 000 live births, 509 women die during the maternal period that spans pregnancy and 6-weeks following pregnancy, ranking Niger with the 20th highest Maternal Mortality Ratio (MMR) globally (20). While progress has been made in reducing maternal mortality in Niger over the past 2 decades, little progress has been made in the recognition and prevention of maternal abuse. There is lack of scientific research to support anecdotal and white/grey literature that has described the existence of abuse of maternal rights, particularly when delivering in the formal healthcare setting in Niger. At the time of writing, there are no published peer-reviewed articles or sources of data on obstetric violence in Niger available via Pubmed, SCOPUS, or Google Scholar.

In this paper we explore women’s experiences of obstetric violence along with specific foundational cultural and systemic elements of maternity care in Niger to inform prevention interventions and training of maternity care providers. The project was designed in accordance with guidance provided by the WHO to reduce the prevalence of obstetric violence by: (1) generating evidence within a health system to promote accountability for the development of policies, and (2) utilizing a rights-based approach to organizing and managing health systems to facilitate the provision of respectful quality care at birth. Our study aimed to establish evidence of the existence of obstetric violence, describe its manifestation, and identify drivers, facilitators and consequences of the abuse of women during labor and delivery in public clinical settings in Niger.

## Methods

### Sample and recruitment

This study uses a qualitative, descriptive phenomenological approach (21) to explore the experiences of women who had given birth at a public health facility in Niamey, Niger, within 5 years from the time of interview. Data was collected from March – August, 2022. The city of Niamey, as are most urban regions, is a mix of multiple ethnicities and many socio-economic levels; therefore, inclusion of participants from various groups was possible. Researchers were initially tasked with conducting 5 key informant interviews (KIIs) with two practitioners, two mothers who were community leaders, and one representative from a non-governmental organization focused on maternal health. Interviews were conducted in hospital offices. Questions focused on experience with, knowledge of, and perceptions of practices towards mothers during pregnancy and delivery. A Listening Session (LS) was conducted with eight mothers recruited through key informants. The LS served to further explore participants’ experiences and perceptions of the treatment of women during maternity care in formal settings. LS is a method of providing a voice to participants and more ethnographic data compared with a structured focus group method. Four general questions guided the LS: (1) What has been your experience with receiving maternity care? (2) What went well during the maternity care you received? (3) What do you wish would have happened better or differently? (4) What recommendations do you have for improving maternity care? LS lasted 75 minutes and took place at the home of a woman who volunteered to host it. The LS was conducted primarily in a mix of Djerma and Hausa languages, based on participants’ comfort. One or both languages were understood by all participants and the two facilitators. The session was recorded and later transcribed directly into French by one of the facilitators, and then verified by another member of the team for accuracy. The LS session provided a general sense of how common obstetric violence might be, guided the development of an individual interview guide, and the identification of strategies for recruiting women for individual, more structured interviews.

Recruitment was purposeful and criterion driven: women having delivered in a public hospital or clinic within the past 5 years, and from the northern, central, and southern region of the metropolitan region of Niamey. We recruited women using snow-ball sampling beginning with LS participants who referred other women to the study. Interested women contacted study staff via telephone or WhatsApp communication platform. Interviews were conducted at the homes, neighborhood or other location identified by the participants. At each interview, following informed consent, participating women provided demographic information including age, years of schooling, occupation, number of pregnancies, number of live children, location of deliveries, prenatal care received, urban/rural residence, marital status, and ethnicity. An important goal of the interviews was to ensure that the women felt their voices were heard and respected. Mothers were asked to share their personal experiences with their last delivery at a public maternity health center: their choice of birthing clinic, their experience during labor and delivery, how they were welcomed by the clinical staff, how the process went, and what the outcomes were. They were also asked about their hopes for maternity care in public health centers in Niger and their suggestions for improving the processes and system.

Individual interviews were conducted with 50 women from three primary areas of the city of Niamey. Interviews were semi-structured, conducted by four women fluent in two to three local languages. Copious notes were taken to record responses to questions and stories of the women. Duration of interviews ranged from 35 to 60 minutes and were conducted in one of three primary languages of Niger: Hausa, spoken by over 80% of the population, Djerma (or Zarma), the local language of the region of Niamey, and French, the official language spoken by approximately 30% of the population. Each participant received compensation of 2,000 CFA francs (approximately 4 USD), an amount proposed by local collaborators and confirmed by key informants. This paper presents collective findings from the key informant interviews, LS, and the individual interviews. Research approval was received from the Issaka Gazobi Maternity Hospital and the University Abdou Moumouni School of Medical Sciences, and the University of Rochester Medical Center Research Subject Review Board.

### Trustworthiness, credibility, and reflexivity

To strengthen trustworthiness and credibility, and to improve rigor of the study findings, several steps were taken (21–23). First, the research team is multidisciplinary (including anthropology, public health, obstetrics and gynecology, and mental health), multinational (comprises both Nigeriens and non-Nigeriens), experts in qualitative methods, stigma and discrimination, clinical care and gender-based violence, and several are multi-lingual (English, French, Hausa and Djerma). Secondly, we used a community-engaged approach to data collection, analysis and interpretation, working with local women community leaders with experience advocating for women’s empowerment and autonomy (24,25). Thirdly, the investigators used bracketing processes to recognize and reduce the impact of their personal, cultural and professional biases and assumptions on data collection and analysis (26). Fourthly, women interviewed had no familial relations the interviewers, and no clinicians conducted interviews.

### Data analysis

Review of KIIs and LS data indicated themes parallel with elements of Bohren et al’s (7) framework of obstetric violence. These elements were used as *a priori* categories to organize the codes and emerging themes from individual interviews. Codes and concepts that were not reflected in the framework were given new labels. Our descriptive phenomenological approach guided our focus on women’s experience while giving birth at a maternity hospital or clinic. We listened to the stories of women with the aim of providing a picture of the treatment of mothers during maternity care and associated socio-structural elements. Responses and notes from interviews were uploaded into Dedoose software and coded line-by-line by one researcher who drafted the initial codebook based on the *a priori* categories, and then discussed with the study team prior to finalizing. Organization of categories and themes were again discussed once all interview data were coded. Discussions allowed for revision of elements from a priori themes and decisions regarding concepts outside of the original framework. KIIs and LS data were used to triangulate data with findings from individual interviews. The analysis was deemed complete once there was 100% agreement between team members. Themes were presented to local women engaged in the research process to discuss and validate interpretation of findings. As this is a descriptive study focused on the phenomenon of birthing, the results are presented as the women described their experiences, while the discussion provides researchers the opportunity to interpret these findings considering the context and existing literature.

## Results

The 58 mothers (8 of which participated in the LS) who shared their stories were between 19 and 40 years of ages (mean age=27), from 4 of the major ethnic groups or the country, primarily homemakers and/or conducted small commerce, and had no schooling or less than 10 years of formal schooling (Table 1). All were or had been married (i.e., divorced). Obstetric histories included 19 miscarriages, 6 infants deceased, an average of 2.9 pregnancies and 2.2 live children. Most (44/58) had had at least one prenatal visit prior to delivery.

**Table 1.** Participating women’s characteristics (N=58).

| Characteristics | Number |
| --- | --- |
| **Age** (range= 19-40; mean= 27) |  |
| **Ethnicity:** Hausa | 22 |
| Zarma/Songhai | 24 |
| Fulani/Bororo | 6 |
| Touareg | 2 |
| Non-Nigerien | 4 |
| **Income Source:** None | 14 |
| Commerce/ business | 15 |
| Small/Small business | 20 |
| Professional | 4 |
| Student/Trainee | 4 |
| **Marital Status:** Married | 56 |
| Divorced | 2 |
| **Schooling:** None | 13 |
| Some elementary/Elementary | 16 |
| Some Secondary | 17 |
| Secondary Diploma/Technical Degree | 3 |
| University/Professional Degree | 9 |
| **Obstetric History:** Currently pregnant | 4 |
| Average number pregnancies | 2.88 |
| Number of miscarriages | 19 |
| Number of children deceased | 6 |
| Average number of children alive | 2.20 |
| Obtained prenatal care | 44 |

The stories of participating women indicate experiences of abuse and disrespect at the hands of maternity hospital or clinic staff, specifically midwives, nurses, and birth-room attendants. The abuses described included elements of all but one (sexual abuse) of the categories of Bohren’s third-order themes of physical and verbal abuse, discrimination and stigma, failure to meet professional standards of care, poor rapport between women and providers, and health systems conditions and constraints. An additional theme in our study is the role of cultural beliefs.

### Cultural Context

Key informants and women interviewed provided important cultural context for their stories. Traditionally, in Niger, a woman’s pain and suffering during childbirth is seen as part of the most important honor, privilege and blessing for women and the best gift for their husbands and families. Consequently, except in extenuating circumstances, women are expected to minimize expressions of pain during birthing as a sign of bravery, similar to a warrior in battle. In fact, the concept of ‘war’ or ‘battle’ emerged from the stories, with women describing birthing as one would a war. Both are honorable because they involve pain and suffering taken for love of family and a sense of duty/responsibility to one’s community; both are honored and expectations of success create added emotional pressure on these highly dangerous tasks; both bring a sense of excitement filled with dread before the event; both have risks of injury or death; and, when there is success, both are rewarded with honor and celebration. The following quotes exemplify these sentiments. *“Giving birth is like going to war: you know you could die,”* and *“Giving birth is between life and death…”*

Key informants explained that prior to a woman giving birth, it is the norm for people to *“visit her in her 8^th^ or 9^th^ month before she has the baby because you want to wish her a good delivery. You don’t know if you are going to see her again. Only Allah knows.”* The expectation is that, despite the pain of giving birth, *“it is important to be courageous.”* When asked to expand on the reasons, some eluded to the potential of attracting evil spirits, and others were not sure: *“that’s just the way it has been. I don’t know. But sometimes you can’t help it…sometimes things go wrong and it’s difficult (to not scream during labor and delivery)”*(mother). One community leader (a grandmother) explained that *“it’s just not good to be screaming and yelling. You have to try to be brave.”*

As these beliefs are upheld and persist, the positive and supportive aspects of giving birth via traditional methods are no longer in existence given the medicalization of maternity care. In the medical context, women are expected to continue to behave bravely regardless of complications or pain resulting from poor medical practices. Traditional protective and supportive elements of birth attendants and support from other women, dignity and respect for privacy, are lacking in clinical settings described by participating women. *“When I was ‘working’ (in labor) this woman was suffering and trying to climb on the [birthing] table and she was having difficulties. She couldn’t do it by herself and the nurses wouldn’t help her. So, I got off of my table and helped her. My ‘work’ (labor) was not yet too far in so I had some strength to help her climb there.”* The women explained that people who accompanied them were not allowed to be with them, and they couldn’t come in the birthing room to see them. All accompaniers would wait outside, on their mats, with the woman’s clothes and baby clothes, for hours until the baby was delivered and the mother was ready to be moved to a recuperation room with other mothers. This isolation from their social support makes them dependent on the maternity hospital workers’ skills, compassion and care. Coupled with the lack of knowledge and/or respect for human rights, lack of adequate training, lack of supervision, low-resources, and traditional beliefs about expressing pain during birthing, women become vulnerable to abuse and poor care.

### Physical abuse

Women interviewed described instances of being slapped in the face, hit in the back of the head and on other parts of their body, and shoved, pulled or pushed. The hitting occurred primarily during labor, on the birthing table, and reasons were mostly because the women expressed pain or did not do as asked by the maternity workers. Shoving, pulling and pushing occurred most often when patients were off the table, usually when they were too slow acting out a command (getting on or off the table, walking to where directed).

*“My water broke during the night; but because I wasn’t able to deliver, the midwife helped me by pushing on my stomach with force to make the baby come out.”*

### Verbal abuse

Common throughout the interviews were staff use of harsh or rude language, judgmental or accusatory comments, threats, and blaming. Expressing pain, asking questions, and non-compliance often led to abuse. Many women described being insulted when they cried out in pain during labor and delivery “*she kept yelling at me saying things like ‘do you think you’re the only woman who has ever given birth? Stop screaming.’*” Another woman was told to stop screaming, “*that’s what you get for spreading your legs*.” Such comments were yelled out or said with a mocking tone. One woman explained, “*I was lucky for my last baby - the doctor was very attentive, but as soon as she left, the nurses and room helpers changed their behaviors towards me and ignored me and then insulted me when I was asking about my baby*.” Another participant stated, *“If you scream, you become subject to mockery. people point fingers. Yet I was suffering so much and couldn’t express my pain in the delivery room.”*

*“I was suffering so much I couldn’t even lift my feet to get on the birthing table. The midwives wouldn’t help me. They told me I was a capricious girl. They treated me like I was a vulgar beggar*.

*It’s another pregnant woman who was also there to give birth who helped me get on the table. She really helped to get over the mockery from the midwives. I will always be grateful to her.”*

### Stigma and Discrimination

Experiences of stigma and discrimination described were based primarily on perceived socio-economic status, which they often immediately judged by the woman’s and her accompanier’s physical appearance (e.g., clothes), ethnicity, mode of transportation to the clinic, (e.g., on foot or public transportation vs. a private vehicle), literacy level, and young age. It is important to note that discrimination was trumped by social network, meaning if someone on the staff knew the woman personally or via other family or friend, she was treated better regardless of her background. One participant explained, “*They were paying more attention to this other woman; I don’t know if they knew her or if it was because she seemed to be well off*.” The difference between the treatment of those the staff knew or who seemed to have more money was repeatedly a concern of the mothers we spoke with. Although most women from families that could afford it went to private clinics where they are treated respectfully. But in the public maternity hospitals and clinics, those who have the least are treated the worst, and it is indeed a battle that they hope to endure and come out alive with their baby. A mother decried, “*I just wish they would treat everyone the same, not just those women they know or the ones that dress nice or look like they have money*.” One mother’s comment summarizes how they felt about being included and listened to, *“I’m so happy that someone is asking about this. I didn’t know what to do, who to tell. I want everyone to know that my baby died because of the midwives and it’s not right.”*

### Failure to meet professional standards of care

Standards of care were not met in the majority of cases and were exemplified through ignoring women and making them wait for long periods of time prior to examining them, performing painful vaginal exams, and hastening delivery by intentional cutting of the vulva and/or painfully pushing the baby out using their elbows. Several mothers explained that they “*had to wait in the hallway or outside*” until they could be seen. Being at the public maternity clinics meant they are at the mercy of the health professionals present. Delays were sometimes because the staff were late, or because they were busy with personal tasks of socializing with other, eating meals, or using their cellular phones for long periods of times. *“They tell us to come very early for consultation. We would come sometimes at 5 o’clock and no one would be there until 7 or 8.” “We can see them talking and having their breakfast together while we were all waiting. We don’t have a choice.”* (LS)

Sometimes nurses or other trained obstetric professionals are absent and oftentimes those present are inadequately trained. “*They said I needed a caesarean but there was no one there who could do it. They took me in an ambulance to another maternity*,” explained a mother. The lack of adequate training was also obvious in the way they described being treated:

“*I was in labor for hours, waiting for someone to pay attention to me. My sister would try to get the nurses to pay attention to me but they ignored her, told her that my baby was obviously not ready, so we should just wait in the hall. She had not even examined me; how would she know? Why didn’t she just check me? I kept crying out that the baby was coming but they would walk by and ignore me. Well, the baby came out and that’s when they finally put us in a room so they can cut the cord and wait for the placenta. I was lucky. The lady next to me was even worse. They took her baby out to wash him and left her bleeding. She died right there with her blood everywhere on the floor. It was traumatizing. I understand women who stay home*.”

“*They killed my baby. It was born but I didn’t hear it cry. They took him out of the room to clean him up and make him cry and I never saw him again. They left me in the recovery room for hours. When I would ask about my baby, they would try to ignore me or say that he was in the other room, that they would get back to me. Hours later, someone finally told me that he had died. I didn’t even get to hold him…I don’t know what happened. No one said anything. I had to go home like that*.”

A young woman described losing her baby and tearing because the nurses tried to force him out of her womb: “*She got on the table and put her elbows on my belly to push the baby out. It was so painful - I can’t describe it.*”

### Poor rapport between women and providers

Communication between providers and women was problematic. Women’s concerns were often dismissed, and staff attitudes demonstrated a lack of empathy and compassion, and neglect, especially towards women that appeared to be unschooled and poor.

*“Giving birth is between life and death, so the midwives should help, show some pity instead of yelling at or scolding patients. They should put themselves in the place of the woman giving birth and feel their pain. They are women. They should understand.”*

However, the few women with university or professional degrees, and those with friends/family members on the staff, described being treated with dignity, respect and kindness, compared with those women in labor at the same time. *“I had a good delivery at the* [redacted] *maternity. All went well because I know the ’majeure’ there. I was even brought home in the hospital vehicle after delivery.”*

*“I felt bad for the others in labor. I was lucky because my mother is friends with the midwife. My mother called to be sure she was working today before bringing me here when*

*I started feeling contractions. She [the nurse] wasn’t here but came to meet us here. That way I could be welcomed and treated well.”*

### Health systems conditions and constraints

Participants and key informants describe the maternity care system as under-resourced in terms of basic materials like gloves and plastic covering for birthing tables, to medical equipment, and poor training of obstetric staff. An important system-level problem described is the selection of staff for training in medical and health fields, which is based on middle school educational tracks that limit options for students. For the most part, maternity care was not a career chosen by the staff. A key informant explained *“The lack of empathy can be understandable. For most of [the clinical staff] they did not choose this work. It’s difficult to do something you did not opt to do or if it is your only option for earning a living.”* Secondly, there are too few obstetricians/gynecologists working in the public field, rendering supervision of clinical staff challenging. Thirdly, the overall economic challenges in a city that has been increasing in living costs while wages have remained unchanged, produced a situation where coercion is not uncommon. When coupled with lack of supervision, this economic situation lends itself to financial coercion on the part of staff towards women in vulnerable situation of delivery. Women described how, despite donations to maternity hospitals advertised in the media claiming to be designed to help low-resourced mothers and infants, women are expected to either come to deliver with all the needed materials, or to purchase them from the clinical staff: *“We know they receive [donations of materials] from NGOs and foreign organization; but they still make us pay for it.”* (LS) Women would be refused care if they did not come prepared with everything, including a plastic bucket with cover for the placenta (which it to be taken home), sheets or cloth for cleaning blood and wrapping the baby, as well as cleaning supplies. Women and the person accompanying them are responsible for removing all soiled cloth and cleaning the space/floors. Similarly, any medication needed during the delivery is to be purchased. Women who had to rush to the clinic and did not have materials with them were refused care until they or their accompanying person purchased the items. They were made to wait outside the labor/delivery room, on the floor in hallways or outside in open air.

*“Women are disoriented because they don’t know where to go and the clinicians don’t give them direction. We’re there, sitting on the floor or on our shoes; there aren’t enough places on the benches when there are many of us.”*(LS)

For those few who explained that they had a short labor and her baby came very fast, they did not escape verbal abuse and financial coercion: *“The baby came before I could get taken to the maternity. We cut the cord and immediately came to the maternity. The midwives were so rude. They said ‘why did you bother coming here? Since you know what to do go back home and finish up on your own, since it looks like you didn’t need us anyway.’”* Another reported: *“They charged me a fine for having the baby at home before coming to the clinic to have the baby checked and weighed. I guess they didn’t make money from the delivery items and they were going to get money from us somehow.”*

### Recommendations for improving safe and respectful maternity care

The women had nowhere to go to express their frustration and report poor treatment. Several mothers echoed this sentiment: “*We want you to tell our stories. We are frustrated and angry but don’t know where to go to report how some nurses treat us at the maternities.*” (LS) When asked what they would suggest to help remedy the issues, women reported simply wanting to be able to give birth without worrying about how they will be treated, and to know what to do when they are mistreated, especially when there are dire consequences like someone dying or losing their baby, or having complications following botched episiotomies and damage to the uterus. Stakeholders interviewed proposed that interventions consider training maternity care staff in respectful delivery of care for pregnant women of all backgrounds, and in the enforcement of these rights through better supervision of maternity staff. *“I don’t see this type of treatment when I’m present. I wasn’t aware that it was that bad in my absence,”* explains an Obstetrician. Having institutional policies and accountability system would help to ensure that women can better trust the system to safely aid their labor and delivery. Many of the women were so traumatized by their treatment and by witnessing the dangerous consequences to women in difficult labor, that they reported being reluctant to return for their next delivery or for reproductive care. Several mentioned if they had to come back, they would try to find a maternity hospital where they might have social connection with the staff. Others planned on delivery at home. One woman’s comment is representative of the overall feeling as she describes her reaction to watching a woman in labor in the shared birthing room bleed to death while the midwives ignored her: *“I will never go back…I will deliver at home, or at my mother’s.”*

## Discussion

To our knowledge, this is the first published study to document women’s experiences of obstetric violence in Niger. The study findings indicate that obstetric violence was a common experience among women who have birth at government run health facilities in Niamey, Niger. Such experiences are a violation of basic human rights and go against the WHO’s Respectful Maternal Care Charter that specifies every woman’s right to: (a) be free from harm and ill treatment; (b) information, informed consent and refusal, and respect for her choices and preferences, including companionship during maternity care; (c) privacy and confidentiality; (d) be treated with dignity and respect; (e) equality, freedom from discrimination, and equitable care; (f) healthcare and to the highest attainable level of health; and (g) liberty, autonomy, self-determination, and freedom from coercion (4).

Study findings corroborate much of the existing literature on women’s experiences of obstetric violence globally and particularly from other African countries. For example, some of the risk factors for the receipt of level of care and respect women received during the delivery process in this study (e.g., judgements made about ethnicity, socio-economic status, place of residence, literacy levels, age, and mode of transportation used to come to the hospital), have also been reported in other studies (3,8–11).

Bohren et al’s (7) typology to categorize and understand women’s experiences during childbirth was very useful to describe women’s experiences in this study (Figure 1). Participants shared many instances of violence and mistreatment during the childbirth process that ranged from experiences of physical abuse, verbal abuse, stigma and discrimination, failure of providers and staff to meet professional standards of care, poor rapport between women and providers, to poor health system conditions and constraints. These findings have been reported in other studies as well (27–29). The systemic issues highlighted in our study are similar to those in similar settings and include lack of resources in hospitals, corruption and financial coercion by clinical staff, poor supervision, and lack of avenues for reporting and enforcing regulations for the protection of maternal rights. These were most pronounced for women of lower socio-economic background, which, in Niger, constitute the majority of the population. The overall poor economy and lack of enforcement of regulations have created an ideal setting for the abuse of power by clinical staff, first by isolating women from their birthing support, and then placing their own financial gain above that of the women they are paid to serve. This is not uncommon in low- and middle-income countries (10).

**Fig. 1.**
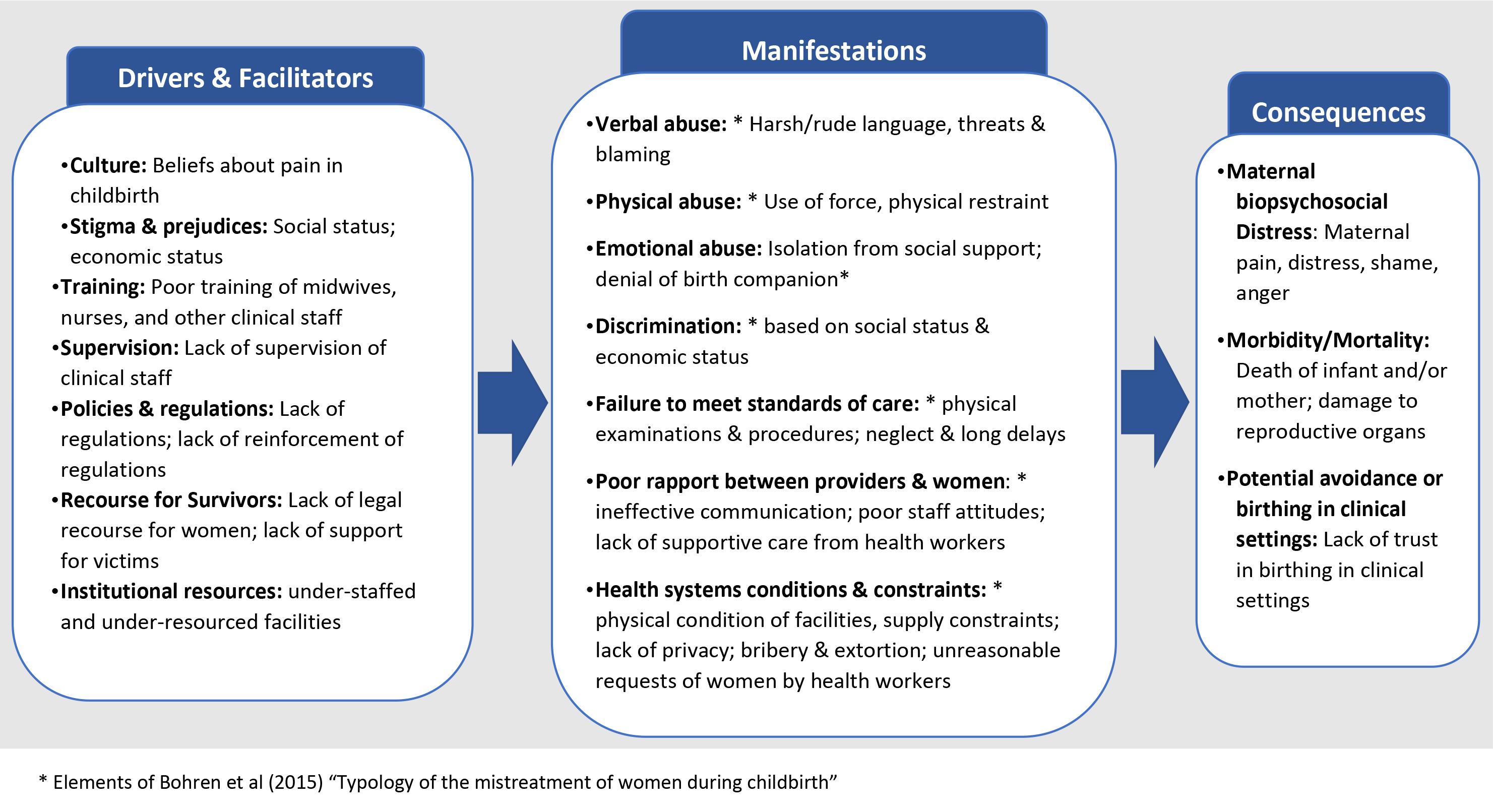
Summary of findings from Niger: Drivers and facilitators, manifestations, and consequences of obstetric violence. This figure presents the list of factors brought up by study participants. Factors are listed by overall category of Drivers and Facilitators (first box), Manifestations (second box) and consequences (third box) of obstetric violence against women. For each sub-category we provide specific elements reported by participants in this study. We use elements of Bohren et al’s (2015) “Typology of the mistreatment of women during childbirth” (7) to describe sub-categories of “manifestations.”

Unique to this study are socio-cultural aspects that were drivers and facilitators of the treatment of women during childbirth at public maternity hospitals and clinics (Figure 1). Firstly, women reported the experienced and observed influence of social networks on childbirth experiences. Participants disclosed that women who were connected with the staff or clinicians at the maternity hospital through friends and/or family, received special treatment that was dignified, respectful, and caring compared to other women who were in labor at the same time, regardless of socio-economic status. While several studies have investigated the association between social networks/capital and facility-based deliveries (30–32), this study highlighted a unique protective function that one’s social connectedness can play in relation to one’s facility-based birth experiences.

Secondly, women’s experiences of obstetric violence in Niger were influenced by cultural beliefs about expression of pain during the birthing process. Several of the participants shared that culturally, braving the pain and suffering of the birthing process is seen as honorable, while expressing pain is a ‘failure’ of a woman’s duty. The process of labor and delivery is viewed similarly to a rite of passage to motherhood, and expressions of pain can attract bad omen for the child and potentially the family. A few other studies have highlighted the impact of specific cultural beliefs regarding pain and traditional practices during childbirth experiences (33–35). For some, pain medication was believed to be harmful to the baby, encouraging them to endure the pain for the sake of their child’s health (36). For others, displaying self-control and not expressing pain during childbirth was considered honorable. Across studies and societies, support from other women or family members during labor and delivery has been valued and serves as important support for enduring pain and suffering (36,37). Beliefs associated with caesarean sections are also explained by cultural beliefs that a woman who does not deliver vaginally has failed and do not deserve the same respect (38–40). Birthing women who received c-sections without understanding the clinical necessity, and without the support and approval of their support system, face internalized stigma and/or experiences of disrespect from their partners, families or communities. In our study, the healthcare workers upheld the beliefs around pain in childbirth but did not consider the difference in context. In public maternity hospitals women are isolated from the traditional support they would receive from other women serving as birth attendants or family members who would help to alleviate the psychological and physical aspects of pain. In fact, accompanying women or partners are not allowed in the birthing room, thereby removing an important aspect of traditional birthing and replacing it with room attendants that tend to multiple women and are not trained appropriately.

Recognizing the importance of traditional birth attendants in the clinical setting, some health systems in Africa have included or called for the inclusion of traditional birth attendants (38,41,42). A study in Ghana described societal beliefs that traditional birth attendants are considered more caring, respectful and better skilled than clinical maternity workers, based on common experiences of poor treatment and poor quality of service at medical facilities (38). The consequences of the maltreatment and poor care of women during labor and delivery consisted of emotional and psychological distress from the disrespect and shame they felt, and the anger of feeling powerless to respond and having no legal or emotional support. Physically, they experienced extreme pain from episiotomies and vaginal tearing resulting from painful procedures, heavy bleeding, infections, and one case of loss of uterus. The extent of morbidity experienced is not measurable, as medical records were not consulted and the women were not aware of the corresponding medical terminology. There is scant research of the extent of the effects of obstetric violence on pregnancy outcomes. One study described evidence of higher risk of post-partum depression among women who experienced abuse during labor and delivery (43). Research from Niger describe incidences of obstetric fistula (44–46), and studies from other countries provide examples of morbidity resulting in part from inadequate maternity care, including infections, damage to reproductive organs, infertility, anemia (43,47). In our study, the ultimate consequence is the death of the mother and/or the infant due to poor treatment, neglect, lack of proper training, and lack of resources.

Given the importance of quality care for better birth outcomes, it is crucial to train clinical staff in safe maternity care and elements of costumer care, especially in countries like Niger where fertility is high, births outside of medical facilities remains, and maternal and infant mortality rates are among the highest in the world.

## Study Strengths and Limitations

This study has several strengths and limitations. The data were collected in partnership with local healthcare centers and key stake holders in the community. We triangulated data from different informants such as providers, women leaders in the community, and women who had delivered a baby within two years of their interview. Multiple languages were used during data collection. The research team was multi-national, multi-linguistic, and multi-disciplinary. Although the primary author and some researchers in the field spoke all four, some words and meaning may have been lost in translation. Also, the findings might be impacted due to recall bias as this is a retrospective study. We tried to minimize the issues with recall by limiting our participants to women who had delivered in the last four years, however, it is possible that women might have experienced some recall-related challenges. Lastly, this study was conducted in Niamey and the findings might not be applicable to other parts of Niger. Nevertheless, as Niamey is the largest city with the largest health system and best geographic access to care, maternity care is expected to be the best in the country. This indicates that the findings could be at either extreme of the spectrum: it represents the worst for those of the lowest income, yet provides access to the best private care for those who can afford it. Our sample was ethnically diverse and representative of the national population.

## Conclusion

Our findings highlight the existence of maltreatment, abuse, and sub-optimal care of women during labor and delivery in clinical settings in Niamey, Niger, in particular for women without economic or social capital. Understanding the cultural context helps to understand meanings, expectations, and practices associated with childbirth (48). Cultural beliefs considered the main facilitators and drivers of obstetric violence in this context include going through the birthing experience with dignity and courage, as exemplified by expressing distress as little as possible, and having a vaginal birth. Fueled by these beliefs, institutional-level conditions and poor training of staff facilitate the manifestation of these abuses of women at their most vulnerable state. Consequently, women experience poor physical and emotional outcomes, higher risk of maternal and newborn mortality, and increased fear of delivering in clinical facilities. Given the high burden of maternal and infant morbidity and mortality in Niger and similar settings, it is crucial that emphasis be placed on training of providers in safe and respectful maternity care, and consider including traditional best practices like the integration of traditional birth attendants in clinical settings to help balance cultural beliefs that may serve as pretext for maltreatment and poor provision of care for birthing women.

## Data Availability

All data produced in the present study are available upon reasonable request to the authors.

## Acknowledgements

Many thanks to the women and practitioners for candidly sharing their experiences with labor and delivery, their reflections and recommendations for improving maternity care in Niger. Special thanks to Association SARA for their partnership in the planning, data collection and interpretation of findings, in particular, Mme. Hadiza Baragé, Mme. Esther Ayouba, and Mme. Roukiatou Sow.

